# Survey on emergency care utilization in tertiary care hospitals in Indonesia

**DOI:** 10.1101/2024.01.18.24301470

**Authors:** Mineto Fujisawa, Kiyomitsu Fukaguchi, Akio Tokita, Yuta Iwamoto, Takanori Takeda, Lukito Condro, Monalisa Tobing, Bobi Prabowo, Rarasanti Rania Qodri, S.Tr. Battra, Tadahiro Goto

**Author notes:** **Correspondence author:** Tadahiro Goto, MD, MPH, PhD, TXP Medical Co. Ltd., Tokyo, Japan; Department of Clinical Epidemiology and Health Economics, School of Public Health, The University of Tokyo, Tokyo, Japan, Address: 7-3-1 Hongo, Bunkyo-ku, Tokyo, 113-0033.

## Abstract

**Objective:** Indonesia’s emergency care system remains suboptimal despite rising demand due to socio-economic changes and increased life expectancy. This study aims to examine patient and family perceptions of the current emergency care system, identify potential needs, and provide a foundation for its development and improvement.

**Methods:** This cross-sectional study used an online survey at two Indonesian hospitals in 2022 to target adult emergency department patients or their families. Each participant’s demographic data, patient journey details, and potential additional medical services were analyzed using descriptive statistics.

**Results:** The study included 446 participants, primarily family members of patients (93.7%), with a median age of 37 (IQR: 28 to 43□years). The majority of patients visited the hospital using private cars (36.9%) and taxis (17.2%), with marked variation between the two hospitals. Only 9.4% of participants were aware of Public Safety Center (PSC) services, and the majority (58.9%) did not use them because they did not know how to contact PSC. The most common waiting time was up to 20 minutes at two tertiary care hospitals. Additional services desired by participants included doctor reservation systems, medical interview applications, and home visitation services. Reported problems included long waiting times and crowded emergency rooms.

**Conclusions:** The study highlights transportation disparities and the demand for supplemental services to better emergency department experiences. For optimizing PSC utilization and future resource distribution, it is vital to further investigate patient behaviors and needs during emergency department visits.

## INTRODUCTION

Indonesia has the fourth largest population in the world. While recent socio-economic changes and increased life expectancy have led to a growing demand for emergency medical services (EMS),^1^ the emergency care system in Indonesia, encompassing EMS and emergency departments (ED), remains suboptimal.^2^ For example, the mortality rate from road traffic accidents per 100,000 population in Indonesia (12.2) was three or four times compared to those in nearby countries such as Singapore (2.8), Australia (5.6) or Japan (4.1).^2^ Moreover, Indonesia’s emergency care system has much room for improvement, as illustrated by the high proportion (62.8%) of patients with myocardial infarction in Indonesia reportedly did not receive timely reperfusion therapy.^3,4^

In an effort to address the rising demand for EMS, the Ministry of Health (MoH) has recently taken the lead in establishing the Public Safety Center (PSC)and an 119 call system in pre-hospital services in Indonesia^5^, and the effectiveness of these measures is expected to increase. A cross-sectional study from a tertiary hospital in Indonesia revealed that ambulance usage accounted for 10% of emergency transportation, while approximately 60% of patients arrived at the ED via private cars and ride-sharing services.^6^ This was largely due to patients and their families being unaware of how to contact an ambulance and the existence of EMS.^5^ However, it remains uncertain whether such findings can be generalized to EDs in other tertiary care hospitals and whether patients and their families are satisfied with the current state of emergency visits and transportation.

Given this context, enhancing Indonesia’s emergency care system is of utmost importance to reduce preventable fatalities and improve public health. The objective of this study is to investigate the perceptions of patients and their families regarding the existing emergency care system, to gain insights into the current state and potential needs in emergency care, and to provide a foundation for the development and improvement of emergency care.

## METHODS

### Study design and setting

This study is a cross-sectional, online survey conducted from January 12th to 24th, 2023 using Google Forms (Alphabet Inc., California, USA). Two hospitals from Indonesia participated in the study: RS Umum Daerah Kabupaten Malang (RSUDKM) and RS Hermina Depok (RSHD).RSUDKM is a governmental tertiary care hospital with 778 employees with 27 ED beds, and RSHD is a private tertiary care hospital with 667 employees with 34 ED beds.

### Study participants

The survey was performed for adult ED patients or their family members who attended the hospital EDs from January 12th to 24th 2023. Surveys were conducted from 10:00 am to 6:00 pm each day of the 13-day study period.

### Contents of the online survey and measurements

This survey consisted of three main sections: 1) respondents’ demographic characteristics, 2) information on the patient’s journey to the hospital, including the patient’s or their family’s prior knowledge, sources of information, means of transportation to the hospital, and satisfaction with emergency medical services, and 3) possibility of additional medical applications to meet potential patient needs. The questions were initially developed in English with a native English speaker, then translated into the local common language, Bahasa Indonesia. The details of questionnaires are shown in **Supplemental Table**. The questionnaire was estimated to take approximately 15 minutes to complete, and the questions were based on a previous study.^6^

### Statistical analysis

The characteristics of the participating hospitals and responders were reported as medians with interquartile ranges (IQRs) for continuous variables and as numbers and percentages (%) for categorical variables. All data analyses were performed using Microsoft Excel version 16.74 (Microsoft Corporation, Redmond, WA, USA).

## RESULTS

### Basic demographic characteristics

The total number of patients who participated in the survey was 446, of which 190 (42.6%) were male (**Table 1**). The median age for all participants was 37□years old (IQR: 28 to 43□years). Among the 446 people who participated in the survey, 418 (93.7%) were family members of patients, 13 (2.9%) were patients themselves and 15 (3.4%) were others. The patients’ homes were most frequently 1-5km away from the hospital (47.0%), followed by 6-10 km (23.0%).

**Table 1.**
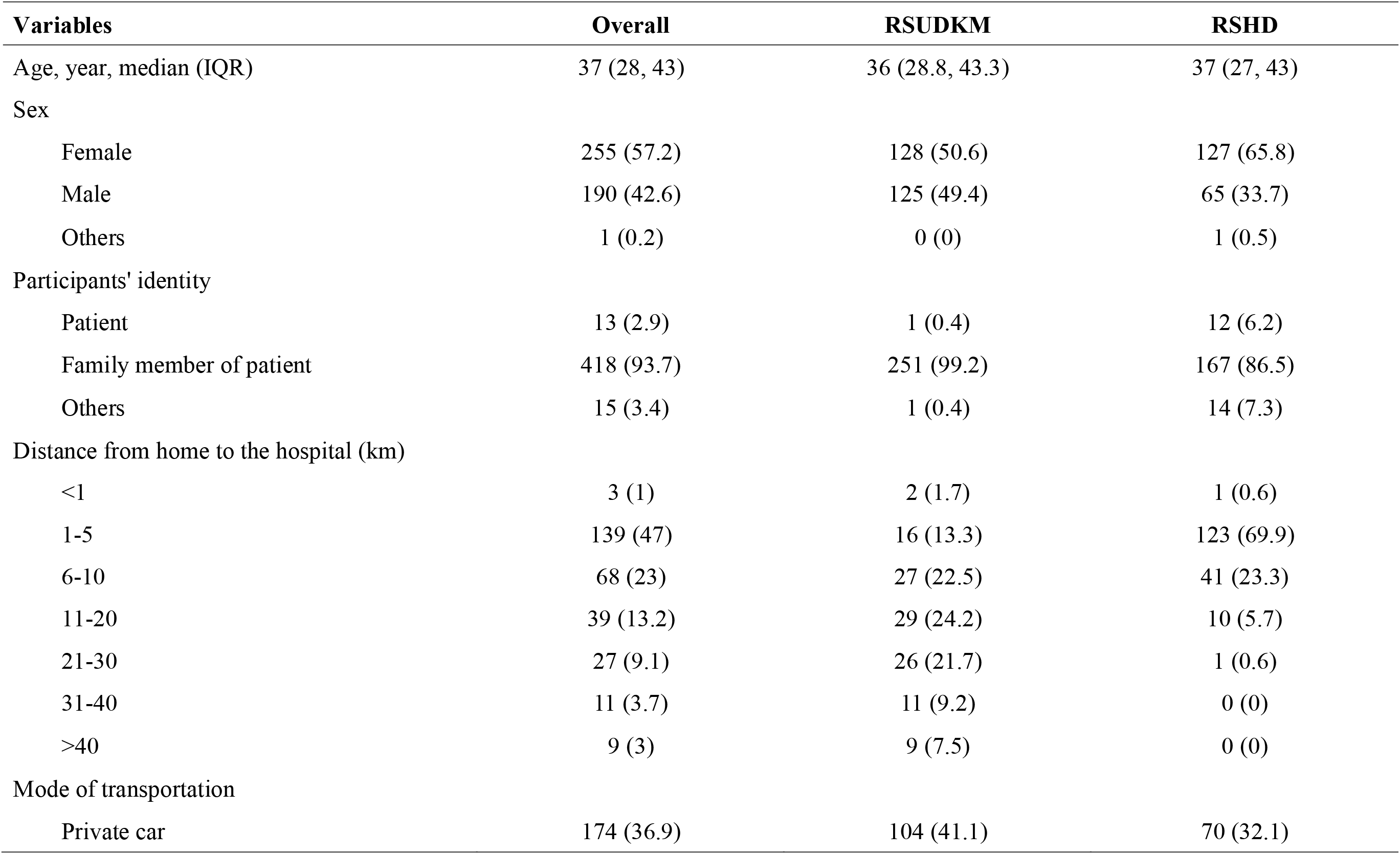
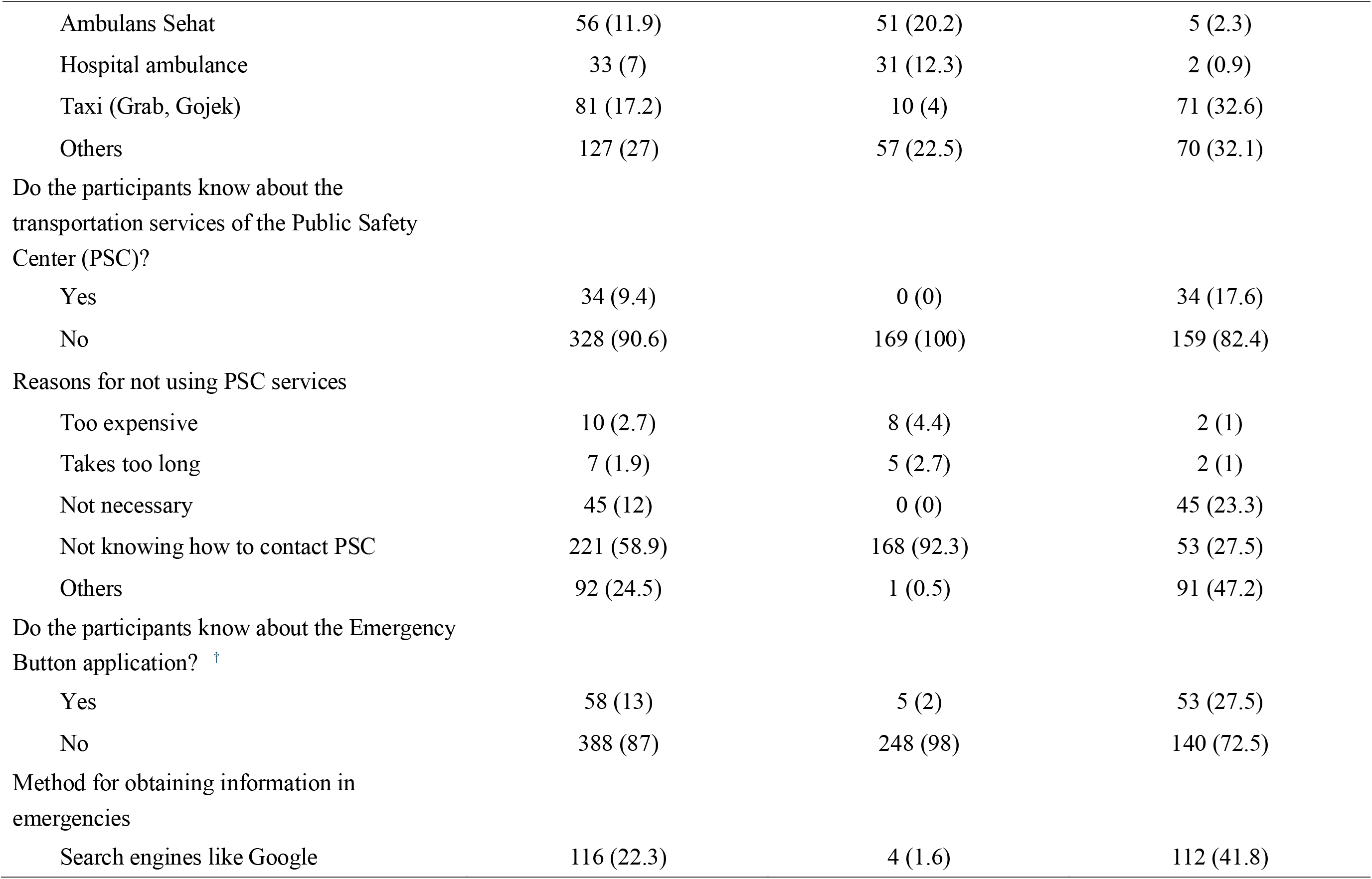
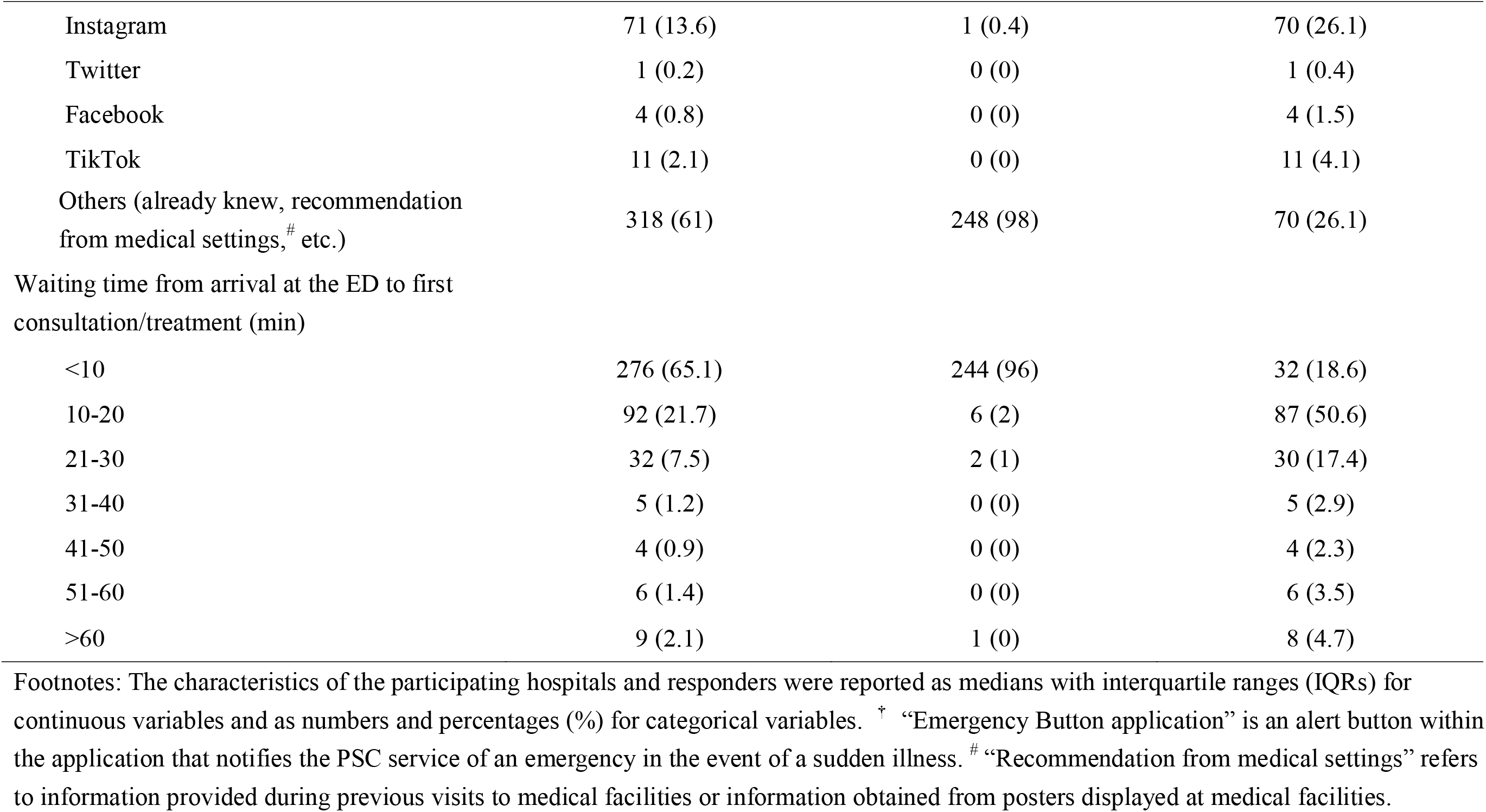
Participants’ characteristics and their responses.

### Transportation modes

**Figure 1A** shows the proportions of transportation modes used for ED visits. Private cars accounted for 36.9% of transportation modes, followed by taxi, including the Indonesian ride-hailing services, Grab and Gojek (17.2%). Ambulans Sehat, a local ambulance service, is responsible for 11.9% of transportations, followed by hospital ambulance at 7.0%. In RSUDKM, compared to RSHD, more patients used Ambulans Sehat (20.2% vs. 2.3%) and Hospital Ambulance (12.3% vs. 0.9%), and fewer used the taxi (4.0% vs. 32.6%).

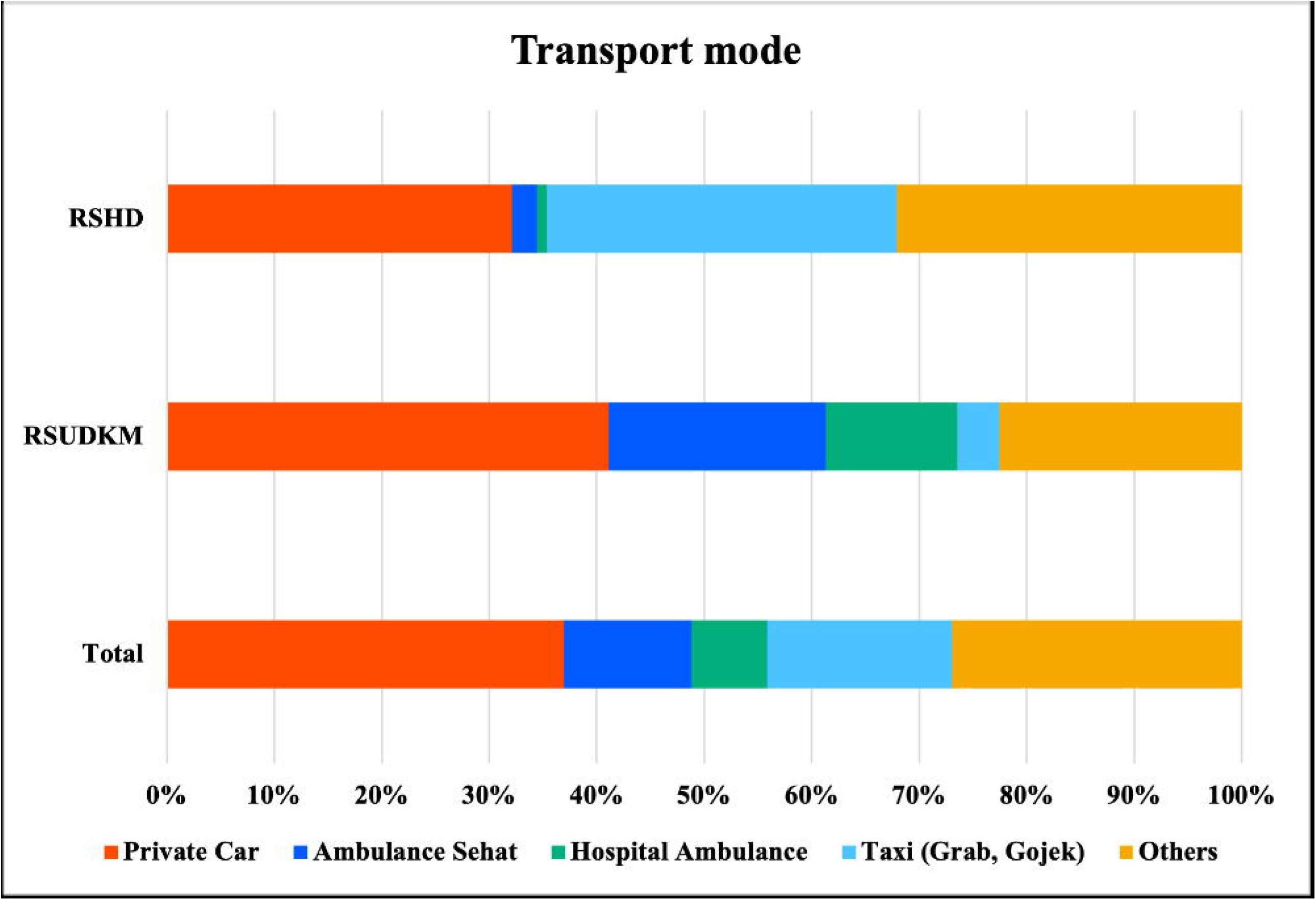

The median score of participants’ satisfaction with the visit was 8 (IQR: 7-9) in both RSUDKM and RSHD. The distributions of satisfaction scores were similar between hospitals. Overall, 9.4% of the participants were aware of PSC services. Furthermore, none of the RSUDKM participants knew about the PSC’s services. **Figure 1B** summarizes the reasons for not using PSC services. The most common reason was that they did not know how to contact the PSC service (58.9%), followed by it being not necessary (12.0%). In the RSHD, only 27.5% of the participants answered that they did not know how to contact the PSC service, while in the RSUDKM, 92.3% of the participants did.

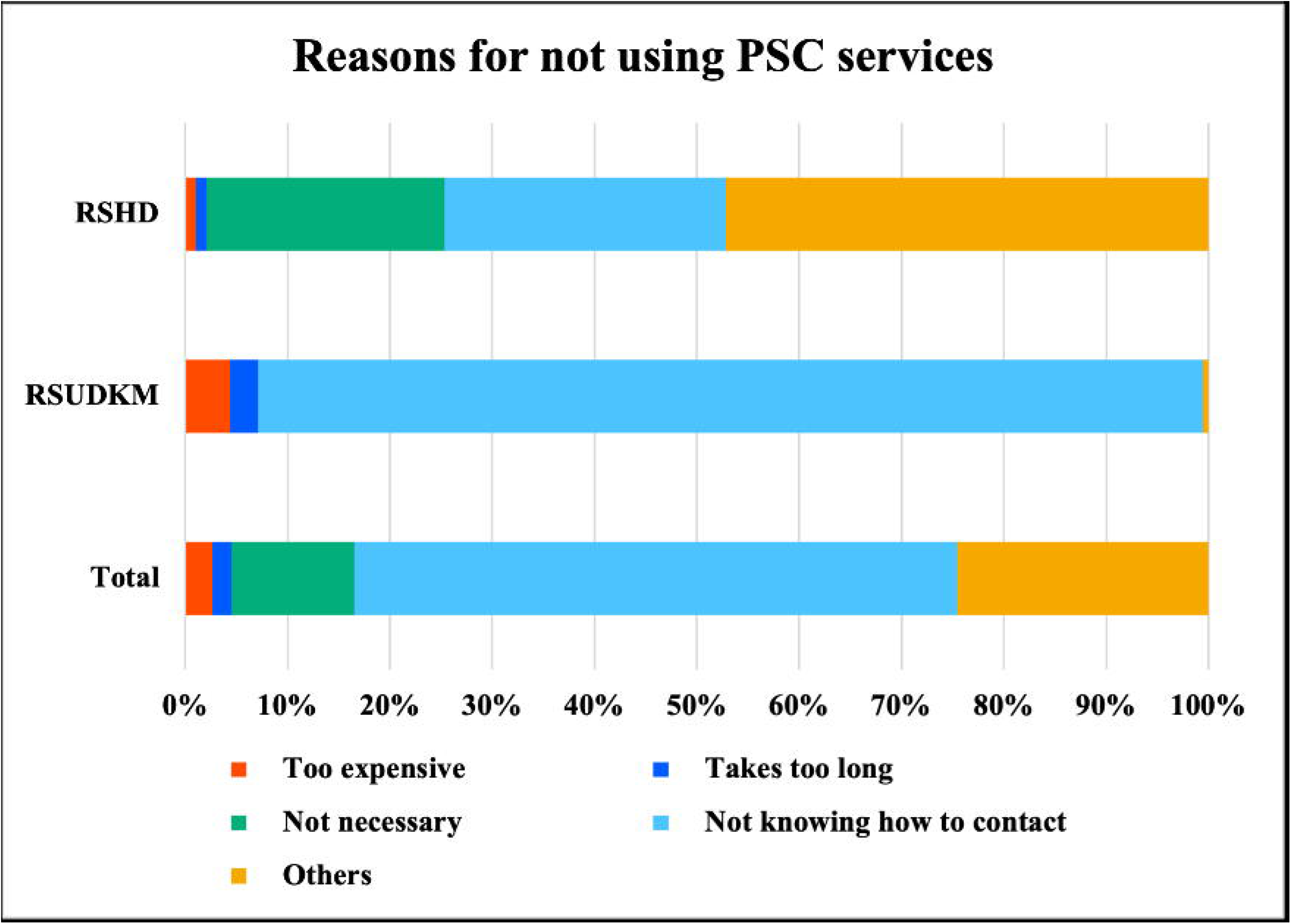

### Patient waiting time

**Figure 2** shows the waiting time from arrival at the ED to the first consultation or treatment. The most common waiting time was less than 10 minutes with RSUDKM and 10-20 minutes with RSHD. Furthermore, 96.0% of all RSUDKM participants had a waiting time of 0 minutes (direct). The median satisfaction level in RSHD was 8 (IQR: 8-9).

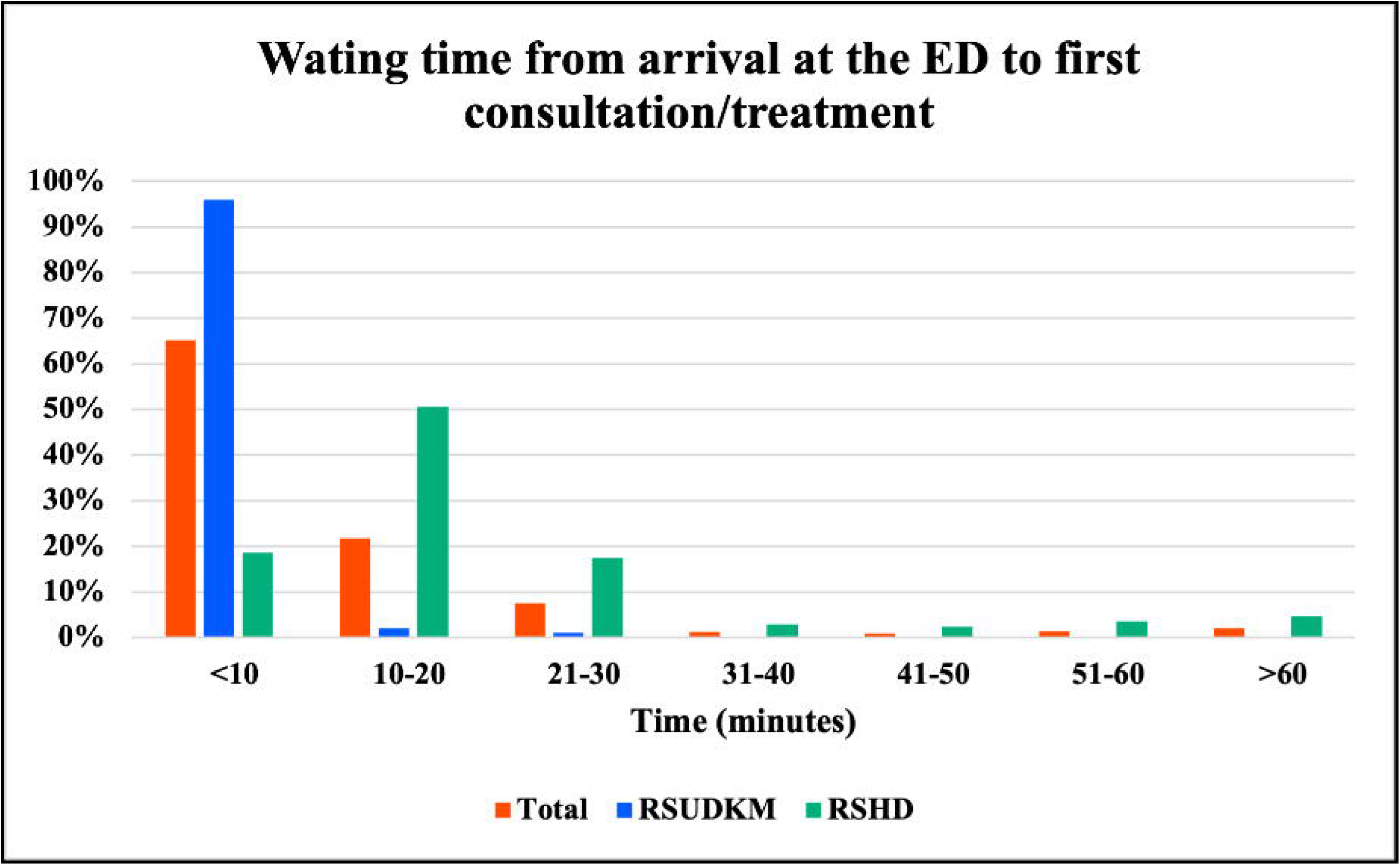

#### Participants’ needs and problems

**Figure 3** shows how much participants were willing to pay for additional medical services in the current emergency system. We focused on the following services: reservation systems to see a doctor, medical interview applications (e.g., smartphone app), and home visitation services. We asked participants if they would like to use each of these services, and 83%, 79%, and 65% of them said they would like to, respectively. For each of the three services, the highest number of respondents wanted to use them free of charge. Apart from “free,” however, the most common price the participants were willing to pay for the three services was Rp 5,000 (0.33 USD) for reservation systems, Rp 10,000 (0.67 USD) for medical interview applications, and Rp 50,000 (3.35 USD) for home visitation services.

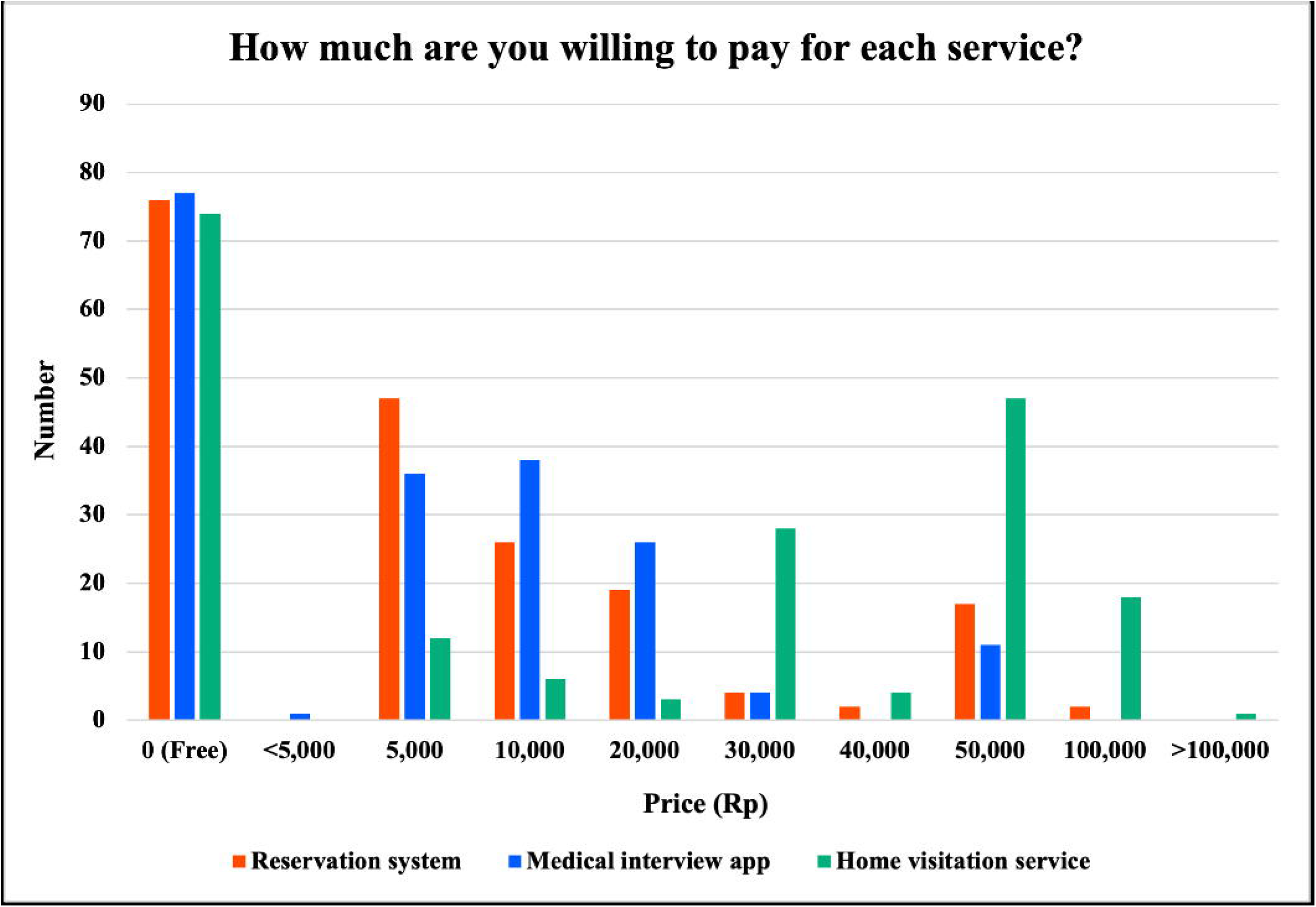

According to an open-ended question on the problems that patients or their families felt in the ER, of the 35 responses received, most participants felt that long waiting times (n=17) and crowded ERs (n=15) were the problems.

## DISCUSSION

In this cross-sectional survey, we found that there was 1) variation in the mode of transportation between hospitals, 2) a shorter ED waiting time than we expected, and 3) a potential need for several additional services, such as reservation systems, medical interview applications, and home visitation services.

In this survey, we found that the most used mode of transportation was private cars, accounting for 36.9%. These findings are consistent with a previous study conducted in Jakarta, Indonesia, which indicated that 30.3% of patients arrived at the hospital by private car.^6^ When Ambulans Sehat and hospital ambulance were combined, ambulances accounted for 18.9%, followed by taxi at 17.2%, with substantial variation (ambulance: 32.5% in RSUDKM vs. 3.2% in RSHD; taxi: 4.0% in RSUDKM vs. 32.6% in RSHD). Such variation in the use of taxi or ambulance has also been reported in earlier studies.^6,7^ For example, in Jakarta, taxi (including ride-sharing services) was the most common method of ED visits (33.5%) while ambulances were used infrequently (9.3%),^6^ whereas 36.7% of ED patients used ambulances in Yogyakarta.^7^

This regional variation in transportation mode can be attributed to several geographic differences.^8-10^ Ride-sharing services are more prevalent in urban areas.^6^ Depok, which is near Jakarta, may have higher usage of such services, which could lead to an increase in taxi usage. In contrast, rural areas tend to have stronger bonds within communities. As a result, patients are more likely to first consult with their local health clinic and then use an ambulance, which is a well-established practice in these communities.

Surprisingly, only 9.4% of participants were aware of PSC, which is the national emergency medical care system. Moreover, approximately 60% of the participants answered that they did not know how to contact PSC as the reason for not using PSC. Of note, none of the participants at RSUDKM knew about PSC, while 18% of the participants at RSHD knew. One possible reason for the low awareness of PSC is that PSC is a relatively new service and has not yet been installed in some areas, and awareness is not fully spread among citizens.^6^ Patients often lack knowledge about the appropriate means of transportation to the hospital, leading to the assumption that private cars or ride-sharing services are commonly utilized. However, in the event of a clearly emergent accident, raising awareness about the availability of Prehospital Emergency Medical Services (PSC) may promote the appropriate utilization of this service.^11^ Another possible reason for the lack of widespread use of ambulances, including PSC, in Indonesia is that ambulances are not given priority on public roads.^6^ It has been pointed out that the ambulance is one of the slowest modes of transportation to the hospital because ambulances cannot run faster than general transportation.

Regarding ED care, the majority of patients (65.1%) experienced a waiting time of 10 minutes or less from their arrival at the ED until the initiation of their first treatment or consultation. Notably, at RSUDKM, 96% of patients had a waiting time of 10 minutes or less. However, it should be noted that in nearly all of these cases, the wait time was actually zero, with patients receiving immediate treatment and consultation upon arrival at the hospital. A study conducted in Jakarta also showed that the waiting time was 5 minutes (IQR 0-10), which aligns with our findings.^6^ These results suggest relatively short waiting times in Indonesian EDs compared to other countries. ^12,13^ Nevertheless, some participants expressed concerns about lengthy waiting periods and overcrowded emergency rooms in this study. Therefore, further investigation is warranted to examine whether long waiting times are a problem in Indonesian EDs.^14,15^ To achieve this, quantitative evaluations through digital modes are preferable.^16,17^

The demand for additional services, namely reservation systems to see a doctor, medical interview applications (e.g., smartphone app), and home visitation services, are high. To address the identified challenges, digitization for these services could be a potential solution, but it remains unclear whether these solutions will actually improve patient convenience, optimization of resources, and finally patient outcomes. Therefore, further research is warranted to ascertain the potential improvements in patient convenience and the optimization of healthcare resources.

## Data Availability

All data produced in the present study are available upon reasonable request to the authors.

## LIMITATIONS

Our study has several limitations. First, not all patients who came to the ED participated in the study. It is possible that a self-selection bias exists in which relatively healthy or health-conscious individuals were more likely to have participated in the survey. However, this bias was more or less mitigated by a dedicated team conducting the survey at fixed times during the study period. Second, this study only collected data from two hospitals in two regions. Since different populations in different regions of Indonesia may have different characteristics, our results may not be representative of the results for Indonesia as a whole.

## CONCLUSIONS

We reported the perceptions of patients and their families regarding the existing emergency care system and potential needs in emergency care, highlighting stark differences in transportation methods and the need for additional services to support emergency department visits. Further investigation is warranted to promote the utilization of PSC and design targeted investigations to improve ED visits and healthcare resource allocation.

