## Supplemental Table for "Survey on emergency care utilization in tertiary care hospitals in Indonesia"

| **Variables** | **Overall** | **RSUDKM** | **RSHD** |
| --- | --- | --- | --- |
| Age, year, median (IQR) | 37 (28, 43) | 36 (28.8, 43.3) | 37 (27, 43) |
| Sex |  |  |  |
| Female | 255 (57.2) | 128 (50.6) | 127 (65.8) |
| Male | 190 (42.6) | 125 (49.4) | 65 (33.7) |
| Others | 1 (0.2) | 0 (0) | 1 (0.5) |
| Participants' identity |  |  |  |
| Patient | 13 (2.9) | 1 (0.4) | 12 (6.2) |
| Family member of patient | 418 (93.7) | 251 (99.2) | 167 (86.5) |
| Others | 15 (3.4) | 1 (0.4) | 14 (7.3) |
| Distance from home to the hospital (km) |  |  |  |
| <1 | 3 (1) | 2 (1.7) | 1 (0.6) |
| 1-5 | 139 (47) | 16 (13.3) | 123 (69.9) |
| 6-10 | 68 (23) | 27 (22.5) | 41 (23.3) |
| 11-20 | 39 (13.2) | 29 (24.2) | 10 (5.7) |
| 21-30 | 27 (9.1) | 26 (21.7) | 1 (0.6) |
| 31-40 | 11 (3.7) | 11 (9.2) | 0 (0) |
| >40 | 9 (3) | 9 (7.5) | 0 (0) |
| Mode of transportation |  |  |  |
| Private car | 174 (36.9) | 104 (41.1) | 70 (32.1) |
| Ambulans Sehat | 56 (11.9) | 51 (20.2) | 5 (2.3) |
| Hospital ambulance | 33 (7) | 31 (12.3) | 2 (0.9) |
| Taxi (Grab, Gojek) | 81 (17.2) | 10 (4) | 71 (32.6) |
| Others | 127 (27) | 57 (22.5) | 70 (32.1) |
| Satisfaction with Transport mode |  |  |  |
| 0 | 0 (0) | 0 (0) | 0 (0) |
| 1 | 0 (0) | 0 (0) | 0 (0) |
| 2 | 0 (0) | 0 (0) | 0 (0) |
| 3 | 1 (0.2) | 1 (0.4) | 0 (0) |
| 4 | 1 (0.2) | 1 (0.4) | 0 (0) |
| 5 | 5 (1.1) | 5 (2) | 0 (0) |
| 6 | 23 (5.2) | 19 (7.5) | 4 (2.1) |
| 7 | 89 (20) | 44 (17.4) | 45 (23.3) |
| 8 | 177 (39.7) | 84 (33.2) | 93 (48.2) |
| 9 | 109 (24.4) | 81 (32) | 28 (14.5) |
| 10 | 41 (9.2) | 18 (7.1) | 23 (11.9) |
| Do the participants know about the transportation services of the Public Safety Center (PSC)? |  |  |  |
| Yes | 34 (9.4) | 0 (0) | 34 (17.6) |
| No | 328 (90.6) | 169 (100) | 159 (82.4) |
| Reasons for not using PSC services |  |  |  |
| Too expensive | 10 (2.7) | 8 (4.4) | 2 (1) |
| Takes too long | 7 (1.9) | 5 (2.7) | 2 (1) |
| Not necessary | 45 (12) | 0 (0) | 45 (23.3) |
| Not knowing how to contact PSC | 221 (58.9) | 168 (92.3) | 53 (27.5) |
| Others | 92 (24.5) | 1 (0.5) | 91 (47.2) |
| Do the participants know about the Emergency Button application? ^†^ |  |  |  |
| Yes | 58 (13) | 5 (2) | 53 (27.5) |
| No | 388 (87) | 248 (98) | 140 (72.5) |
| To what extent would the participants recommend the Emergency Button application to family and friends? |  |  |  |
| 0 | 1 (0.2) | 0 (0) | 1 (0.5) |
| 1 | 0 (0) | 0 (0) | 0 (0) |
| 2 | 0 (0) | 0 (0) | 0 (0) |
| 3 | 0 (0) | 0 (0) | 0 (0) |
| 4 | 1 (0.2) | 1 (0.4) | 0 (0) |
| 5 | 6 (1.3) | 2 (0.8) | 4 (2.1) |
| 6 | 71 (15.9) | 42 (16.6) | 29 (15) |
| 7 | 166 (37.2) | 98 (38.7) | 68 (35.2) |
| 8 | 123 (27.6) | 68 (26.9) | 55 (28.5) |
| 9 | 40 (9) | 27 (10.7) | 13 (6.7) |
| 10 | 38 (8.5) | 15 (5.9) | 23 (11.9) |
| Method for obtaining information in emergencies |  |  |  |
| Search engines like Google | 116 (22.3) | 4 (1.6) | 112 (41.8) |
| Instagram | 71 (13.6) | 1 (0.4) | 70 (26.1) |
| Twitter | 1 (0.2) | 0 (0) | 1 (0.4) |
| Facebook | 4 (0.8) | 0 (0) | 4 (1.5) |
| TikTok | 11 (2.1) | 0 (0) | 11 (4.1) |
| Others (already knew, recommendation from medical settings,^#^ etc.) | 318 (61) | 248 (98) | 70 (26.1) |
| Did the patients search for a hospital via the Internet? |  |  |  |
| Yes | 52 (26.9) | - | 52 (26.9) |
| No | 141 (73.1) | - | 141 (73.1) |
| Waiting time from arrival at the ED to first consultation/treatment (min) |  |  |  |
| <10 | 276 (65.1) | 244 (96) | 32 (18.6) |
| 10-20 | 92 (21.7) | 6 (2) | 87 (50.6) |
| 21-30 | 32 (7.5) | 2 (1) | 30 (17.4) |
| 31-40 | 5 (1.2) | 0 (0) | 5 (2.9) |
| 41-50 | 4 (0.9) | 0 (0) | 4 (2.3) |
| 51-60 | 6 (1.4) | 0 (0) | 6 (3.5) |
| >60 | 9 (2.1) | 1 (0) | 8 (4.7) |
| If there was a reservation system for medical consultations, would the patients use it? |  |  |  |
| Yes | 160 (82.9) | - | 160 (82.9) |
| No | 33 (17.1) | - | 33 (17.1) |
| How much are the patients willing to pay each time they use the reservation system? (Rp) |  |  |  |
| 0 (Free) | 76 (39.4) | - | 76 (39.4) |
| <5,000 | 0 (0) | - | 0 (0) |
| 5,000 | 47 (24.4) | - | 47 (24.4) |
| 10,000 | 26 (13.5) | - | 26 (13.5) |
| 20,000 | 19 (9.8) | - | 19 (9.8) |
| 30,000 | 4 (2.1) | - | 4 (2.1) |
| 40,000 | 2 (1) | - | 2 (1) |
| 50,000 | 17 (8.8) | - | 17 (8.8) |
| 100,000 | 2 (1) | - | 2 (1) |
| >100,000 | 0 (0) | - | 0 (0) |
| If there was a medical interview app that could reduce waiting time at the hospital, would participants use it? |  |  |  |
| Yes | 402 (90.1) | 249 (98.4) | 153 (79.3) |
| No | 44 (9.9) | 4 (1.6) | 40 (20.7) |
| How much are the patients willing to pay each time they use the medical interview app? (Rp) |  |  |  |
| 0 (Free) | 77 (39.9) | - | 77 (39.9) |
| <5,000 | 1 (0.5) | - | 1 (0.5) |
| 5,000 | 36 (18.7) | - | 36 (18.7) |
| 10,000 | 38 (19.7) | - | 38 (19.7) |
| 20,000 | 26 (13.5) | - | 26 (13.5) |
| 30,000 | 4 (2.1) | - | 4 (2.1) |
| 40,000 | 0 (0) | - | 0 (0) |
| 50,000 | 11 (5.7) | - | 11 (5.7) |
| 100,000 | 0 (0) | - | 0 (0) |
| >100,000 | 0 (0) | - | 0 (0) |
| Would you use a private home visit service where a doctor or nurse comes directly to your home instead of going to the hospital? |  |  |  |
| Yes | 124 (64.6) | - | 124 (64.6) |
| No | 68 (35.4) | - | 68 (35.4) |
| How much are the patients willing to pay each time they use the private home visit service, apart from the medical consultation fee? (Rp) |  |  |  |
| 0 (Free) | 74 (38.3) | - | 74 (38.3) |
| <5,000 | 0 (0) | - | 0 (0) |
| 5,000 | 12 (6.2) | - | 12 (6.2) |
| 10,000 | 6 (3.1) | - | 6 (3.1) |
| 20,000 | 3 (1.6) | - | 3 (1.6) |
| 30,000 | 28 (14.5) | - | 28 (14.5) |
| 40,000 | 4 (2.1) | - | 4 (2.1) |
| 50,000 | 47 (24.4) | - | 47 (24.4) |
| 100,000 | 18 (9.3) | - | 18 (9.3) |
| >100,000 | 1 (0.5) | - | 1 (0.5) |
| Satisfaction with overall experience from the moment the patients decided to visit the ER until therapy ended |  |  |  |
| 0 | 0 (0) | - | 0 (0) |
| 1 | 0 (0) | - | 0 (0) |
| 2 | 0 (0) | - | 0 (0) |
| 3 | 0 (0) | - | 0 (0) |
| 4 | 0 (0) | - | 0 (0) |
| 5 | 2 (1) | - | 2 (1) |
| 6 | 5 (2.6) | - | 5 (2.6) |
| 7 | 37 (19.2) | - | 37 (19.2) |
| 8 | 87 (45.1) | - | 87 (45.1) |
| 9 | 28 (14.5) | - | 28 (14.5) |
| 10 | 34 (17.6) | - | 34 (17.6) |
| Satisfaction with call center staff response |  |  |  |
| 0 | 0 (0) | - | 0 (0) |
| 1 | 0 (0) | - | 0 (0) |
| 2 | 0 (0) | - | 0 (0) |
| 3 | 0 (0) | - | 0 (0) |
| 4 | 0 (0) | - | 0 (0) |
| 5 | 0 (0) | - | 0 (0) |
| 6 | 0 (0) | - | 0 (0) |
| 7 | 0 (0) | - | 0 (0) |
| 8 | 3 (50) | - | 3 (50) |
| 9 | 0 (0) | - | 0 (0) |
| 10 | 3 (50) | - | 3 (50) |
| Satisfaction with time until the ambulance arrived and then got to the hospital |  |  |  |
| 0 | 0 (0) | - | 0 (0) |
| 1 | 0 (0) | - | 0 (0) |
| 2 | 0 (0) | - | 0 (0) |
| 3 | 0 (0) | - | 0 (0) |
| 4 | 0 (0) | - | 0 (0) |
| 5 | 0 (0) | - | 0 (0) |
| 6 | 0 (0) | - | 0 (0) |
| 7 | 0 (0) | - | 0 (0) |
| 8 | 4 (66.7) | - | 4 (66.7) |
| 9 | 0 (0) | - | 0 (0) |
| 10 | 2 (33.3) | - | 2 (33.3) |
| Satisfaction with first aid and field response from paramedics |  |  |  |
| 0 | 0 (0) | - | 0 (0) |
| 1 | 0 (0) | - | 0 (0) |
| 2 | 0 (0) | - | 0 (0) |
| 3 | 0 (0) | - | 0 (0) |
| 4 | 0 (0) | - | 0 (0) |
| 5 | 0 (0) | - | 0 (0) |
| 6 | 0 (0) | - | 0 (0) |
| 7 | 0 (0) | - | 0 (0) |
| 8 | 4 (66.7) | - | 4 (66.7) |
| 9 | 0 (0) | - | 0 (0) |
| 10 | 2 (33.3) | - | 2 (33.3) |
| Satisfaction with experience while waiting in the ER from arrival at the hospital until treatment began |  |  |  |
| 0 | 0 (0) | - | 0 (0) |
| 1 | 0 (0) | - | 0 (0) |
| 2 | 0 (0) | - | 0 (0) |
| 3 | 0 (0) | - | 0 (0) |
| 4 | 0 (0) | - | 0 (0) |
| 5 | 2 (1) | - | 2 (1) |
| 6 | 7 (3.6) | - | 7 (3.6) |
| 7 | 39 (20.2) | - | 39 (20.2) |
| 8 | 93 (48.2) | - | 93 (48.2) |
| 9 | 30 (15.5) | - | 30 (15.5) |
| 10 | 22 (11.4) | - | 22 (11.4) |
| Satisfaction with medical treatment and examination |  |  |  |
| 0 | 0 (0) | - | 0 (0) |
| 1 | 0 (0) | - | 0 (0) |
| 2 | 0 (0) | - | 0 (0) |
| 3 | 0 (0) | - | 0 (0) |
| 4 | 0 (0) | - | 0 (0) |
| 5 | 1 (0.5) | - | 1 (0.5) |
| 6 | 1 (0.5) | - | 1 (0.5) |
| 7 | 28 (14.5) | - | 28 (14.5) |
| 8 | 93 (48.2) | - | 93 (48.2) |
| 9 | 46 (23.8) | - | 46 (23.8) |
| 10 | 24 (12.4) | - | 24 (12.4) |
| Satisfaction with experience while waiting in the ER from completion of medical treatment to discharge or hospitalization |  |  |  |
| 0 | 0 (0) | - | 0 (0) |
| 1 | 0 (0) | - | 0 (0) |
| 2 | 0 (0) | - | 0 (0) |
| 3 | 0 (0) | - | 0 (0) |
| 4 | 0 (0) | - | 0 (0) |
| 5 | 1 (0.5) | - | 1 (0.5) |
| 6 | 3 (1.6) | - | 3 (1.6) |
| 7 | 27 (14.1) | - | 27 (14.1) |
| 8 | 93 (48.4) | - | 93 (48.4) |
| 9 | 46 (24) | - | 46 (24) |
| 10 | 22 (11.5) | - | 22 (11.5) |
| Particular problems/obstacles in the ED |  |  |  |
| Long waiting time | 17 (48.6) | - | 17 (48.6) |
| Crowded ER | 15 (42.9) | - | 15 (42.9) |
| Doctors seldom come | 1 (2.9) | - | 1 (2.9) |
| Unfriendly | 1 (2.9) | - | 1 (2.9) |
| Slow response | 1 (2.9) | - | 1 (2.9) |

Footnotes: The characteristics of the participating hospitals and responders were reported as medians with interquartile ranges (IQRs) for continuous variables and as numbers and percentages (%) for categorical variables. ^†^ “Emergency Button application” is an alert button within the application that notifies the PSC service of an emergency in the event of a sudden illness. ^#^ “Recommendation from medical settings” refers to information provided during previous visits to medical facilities or information obtained from posters displayed at medical facilities.
